# Potentially inappropriate prescribing and falls-risk increasing drugs in people who have experienced a fall; a systematic review and meta-analysis

**DOI:** 10.1101/2025.04.07.25325256

**Authors:** Tim O’Reilly, Jessica Gómez Lemus, Laura Booth, Barbara Clyne, Caroline McCarthy, Kinda Ibrahim, Wade Thompson, Christine McAuliffe, Frank Moriarty

## Abstract

**Background:** As certain medications increase risk of falls, it is important to review and optimise prescribing in those who have fallen to reduce risk of recurrent falls.

**Objectives:** To systematically review evidence on the prevalence and types of potentially inappropriate prescribing (PIP), including falls-risk increasing drug (FRID) use, in fallers.

**Methods:** A systematic search was conducted in July 2024 in MEDLINE, EMBASE, CINAHL, and Google Scholar using keywords for fall events, inappropriate prescribing, and FRIDs. Observational studies (cohort, case-control, cross-sectional, before-after) and randomised trials were included. Studies were eligible where participants had experienced a fall and PIP (including FRID use) was reported. Random-effects meta-analyses were conducted to pool prevalence of inappropriate prescribing and mean number of inappropriate prescriptions across studies, with stratified analysis to assess heterogeneity.

**Results:** Fifty papers reporting 46 studies met the inclusion criteria. All studies assessed FRIDs, and twenty-nine assessed other PIP. The prevalence of PIP at the time of the fall was reported in 43 studies, and the pooled estimate was 68.6% (95%CI 66.1-71.2%). Among 23 studies reporting it, the mean number of inappropriate prescriptions per participant was 2.21 (95%CI 1.98-2.45). The most common FRIDs prescribed were sedatives/hypnotics, opioids, diuretics, and antidepressants. Twenty-one studies assessed changes in PIP prevalence post-fall; nine reported decreasing prevalence, with others noting increases/no change/mixed results.

**Conclusion:** Inappropriate prescribing is highly prevalent among fallers, with cardiovascular and psychotropic drugs being the most common. This suggests significant scope to optimise medicines use in these patients to potentially reduce falls risk and improve outcomes.

## Introduction

Medication-related harm is a growing concern and international priority in improving patient safety.(1) Falls are a common adverse outcome which may have medication-related contributors,(2–4) and are often recurrent. Falls can have a significant and often profound impact on people who experience them, such as reduced mobility or independence, premature admission into long-term care, and negative impacts on mental health.(5–7) A fall may present an important opportunity to optimise medication, address potentially inappropriate prescribing or PIP (i.e. posing more risks than benefits for a patient), and reduce the risk of future falls and fractures, or other medication-related adverse outcomes.(8, 9) As falls are multifactorial, current guidelines recommend that medicines review forms part of multi-faceted interventions to reduce falls risk.(10) Adverse events such as falls can prompt reactive medicines review and deprescribing, which occur less frequently in the absence of such triggering events.(11)

Understanding the scope for medicines optimisation among fallers is important to inform targeting of interventions, including which types of drugs are most often implicated in PIP for this group. A previous systematic review quantified the use of falls-risk increasing drugs (FRIDs) among older adults with a fall-related injury and identified 14 studies where prevalence exceeded 65% and no reduction in prevalence was seen after a healthcare encounter.(12) However, this review did not consider other non-injurious falls, nor examine other aspects of potentially inappropriate prescribing, not relevant to falls, and where broader medicines optimisation could be targeted among fallers to improve patient outcomes.

Therefore, the overall aim of this systematic review is to investigate the prevalence of PIP (including the use of FRIDs) in people with a fall or fall-related injury/event. A secondary aim is to determine the types of drugs most often implicated, and whether the prevalence of PIP changes after a fall.

## Methods

This systematic review was pre-registered on PROSPERO (CRD42023417534), conducted in line with JBI guidance,(13) and is reported according to the Preferred Reporting Items for Systematic reviews and Meta-Analyses (PRISMA) statement.(14)

### Eligibility criteria

The following eligibility criteria were applied:

**Study type:** We included observational studies (cohort, before-after, case-control, cross sectional), and randomised trials where the population studied were people who had fallen. Systematic reviews were excluded however any relevant reviews were examined for potentially eligible studies. Other publication types (e.g. conference abstracts, study protocols, commentaries, case series) were excluded. **Population:** We included studies focusing on adults of any age who experienced a fall (based on any definition). Fall-related events such as fracture and syncope were also included. Studies in which the sample population was people attending a falls clinic or similar were included where 70% or above of study participants had a fall or a fracture or where less than 70% of participants had a fall or fracture but characteristics of fallers such as age, sex and prevalence of PIP were reported. For case-control studies, only those studies with falls as an outcome that assessed medication exposure within 90 days prior to the fall (indicating likely medication use at the time of fall) were included. Studies were excluded where people who had fallen were an incidental subgroup of the main study population, and not part of the study inclusion criteria.

**Outcome:** We included studies reporting prevalence of PIP, defined using any approach (e.g. validated tools, lists of medication, specific criteria or indicators, local definitions or where medication use was implicitly judged to be inappropriate). This included any FRID use (as continuing FRIDs in fallers was considered potentially inappropriate), other inappropriate prescribing that is deemed to be relevant to fallers (e.g. anticoagulant use due to increased likelihood of bleeding with recurrent falls, omission of bone protection treatment) and any inappropriate prescribing unrelated to falls (including inappropriate omissions). Studies that assessed inappropriate prescribing of only single drugs/drug classes were excluded.

### Information sources and search strategy

MEDLINE (ovid), EMBASE, CINAHL and Google scholar (via Harzing’s Publish or Perish)(15) were searched from inception up to the search date of 5^th^ July 2024. The search strategy (included in supplementary tables 1-4) was developed using a combination of subject headings, keywords and synonyms relating to PIP or FRIDs and falls or fractures. No language or other restrictions were used. Any results which were in a different language were translated using online translation software Deepl (www.deepl.com). Grey literature sources were not searched, as given the subject it was anticipated that such sources would be unlikely to contribute significantly.

### Selection process

Results from each database were combined, and one reviewer deduplicated using Endnote. Remaining results were uploaded to Rayyan, and its deduplication function was used. Pilot title and abstract screening was conducted on 50 records in Rayyan to ensure the eligibility criteria were applied consistently by all reviewers. Following this, each title and abstract was screened independently by two reviewers (of TOR, LB, JGL, FM). Any disagreements were discussed to reach consensus, or failing this, a third independent reviewer (of CMcA, FM, TOR) assessed the study. A similar process was followed for full-texts, with a pilot on three studies followed by independent review in duplicate, with disagreements resolved as before.

### Data Extraction and Data Items

A Microsoft Excel sheet was developed to extract data and piloted using three studies. Data was extracted by two independent reviewers (of TOR, LB, JGL). Once complete, a third reviewer (FM) checked data for consistency and accuracy across reviewers and studies. Data was extracted on:

- **Study Information:** Design, sample size, time frame, geographical location, setting, duration of follow up.
- **Participant information:** Demographics, definition of falls used, proportion with fall/fracture or where reported, proportion of distinct types of falls/fractures.
- **Outcome information**: definitions of PIP (including FRID use categorised as psychotropic, cardiovascular, and other classes),(2–4) time frame prevalence was measured over, prevalence of PIP, prevalence of specific drug classes (involved in PIP/FRID use), and any change in prevalence post-fall.

### Outcomes

The primary outcome measure was prevalence or mean number of PIP among fallers. For studies reporting prevalence at multiple time points, the time point closest to the fall was recorded, likely reflecting the medications being taken at the time of the fall. For before-and-after studies and randomised trials, if the prevalence of inappropriate prescribing was reported after a fall, unless explicitly stated that no medication changes had occurred, the prevalence at the latest pre-fall time point was recorded. The change in prevalence of PIP at later time points post-fall, where reported, was extracted as a secondary outcome.

### Quality assessment

Study quality was evaluated using the JBI Prevalence Critical Appraisal tool.(16) This assessment was conducted independently by two reviewers (of TOR, LB, JGL) during the data extraction process.

### Data Analysis

Characteristics of included studies were summarised descriptively. Meta-analysis of prevalence estimates, and mean number of PIP was performed using the *metan* package in Stata.(17) Heterogeneity can be a concern in meta-analysis of prevalence; the I^2^ statistic and Cochran’s Q test are reported despite limitations,(18) and this was supplemented by stratifying analyses by study characteristics (i.e. setting, inclusion of fractures, time frame for prevalence measurement, and PIP definition).

## Results

From 3,909 records identified, after deduplication 2,789 titles/abstracts were screened, and 164 publications underwent full-text review (see Figure 1).(19) Overall, 50 publications, relating to 46 studies, met the eligibility criteria. Two studies reported relevant results in two publications each, (20–23) while another study had its results published across three different publications.(24–26)

**Figure 1.**
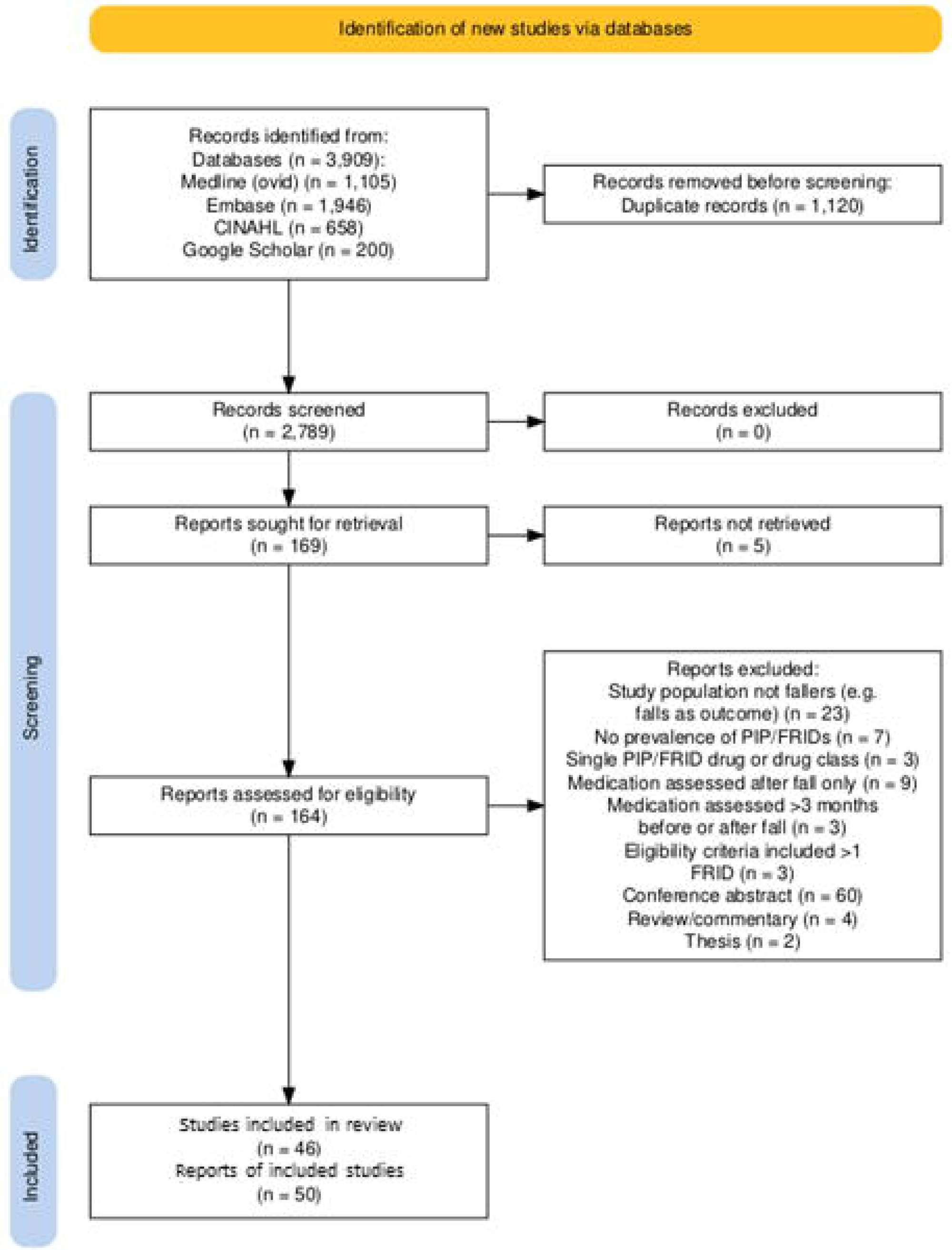
PROSPERO flow diagram of included/excluded publications.

### Quality assessment

Included studies were mostly of high quality (supplementary table 5), however twenty studies had small sample sizes (n<200), while nine studies did not describe the study setting in detail.

### Study characteristics

Of the 46 studies published between 2007 and 2024 (Table 1), nineteen were cohort studies, eleven were cross-sectional studies, ten were case-control studies, four were before-and-after studies, one was a quasi-experimental study, and one was a randomised controlled trial. Studies were performed in the USA (n=9, one conducted in the USA/Mexico border region), Japan (n=5), Sweden (n=4), France (n=3), United Kingdom (n=3), Australia (n=2), Czech Republic (n=2), Ireland (n=2), Netherlands (n=2), Taiwan (n=2), with one study each from Austria, Belgium, Brazil, Canada, Colombia, Denmark, Finland, Germany, Malaysia, New Zealand, Norway, and Spain.

**Table 1.**
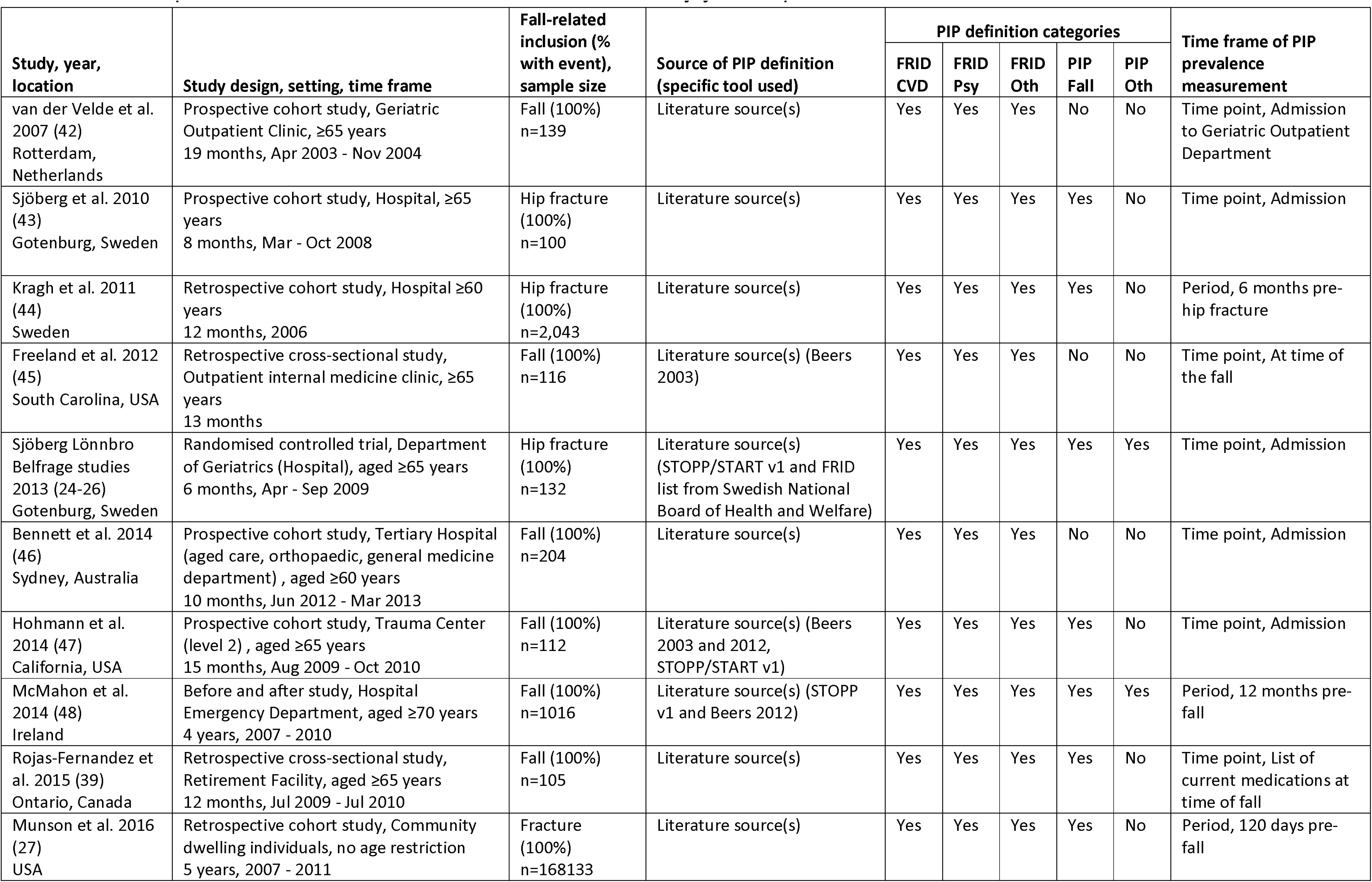
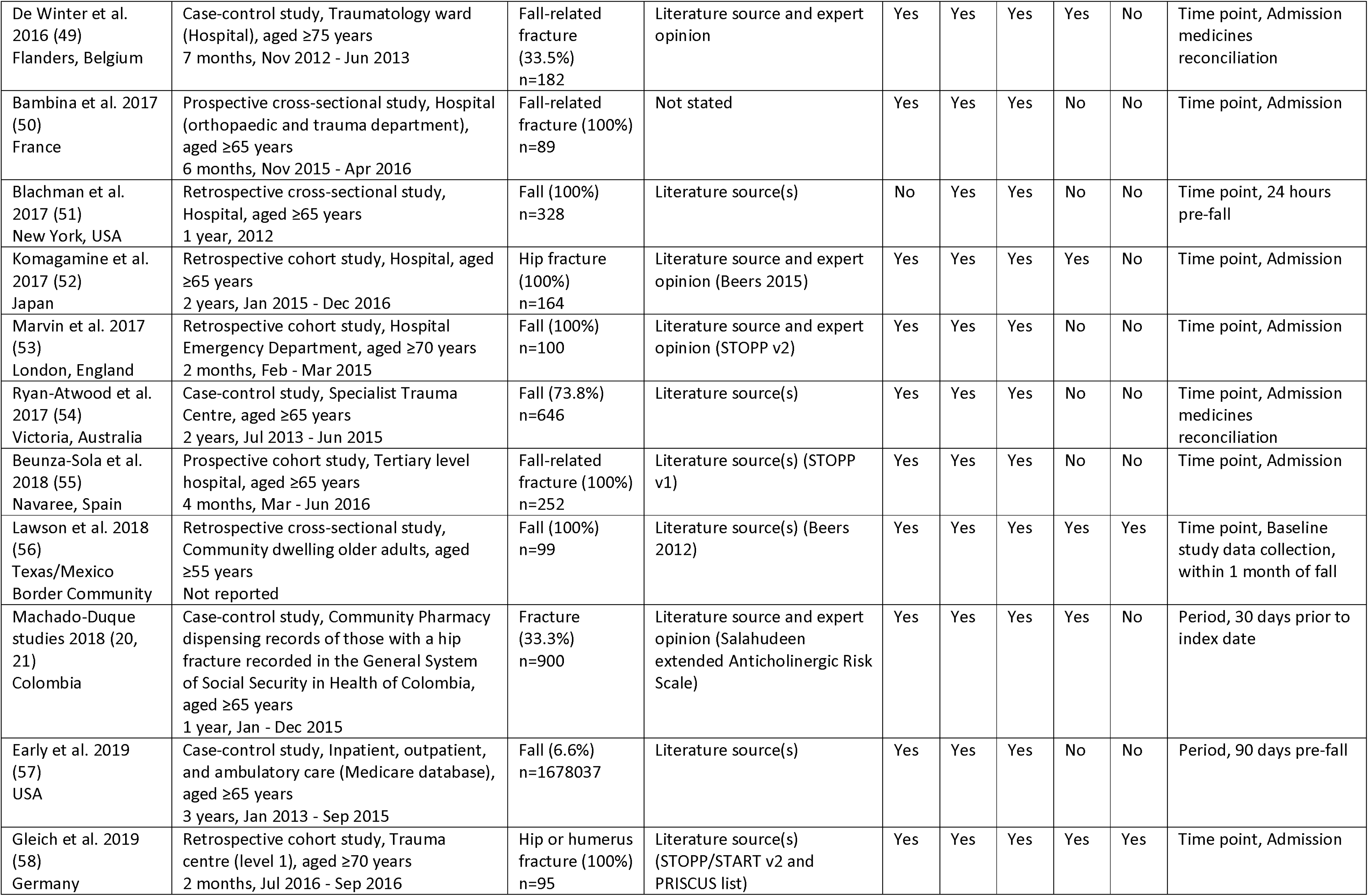
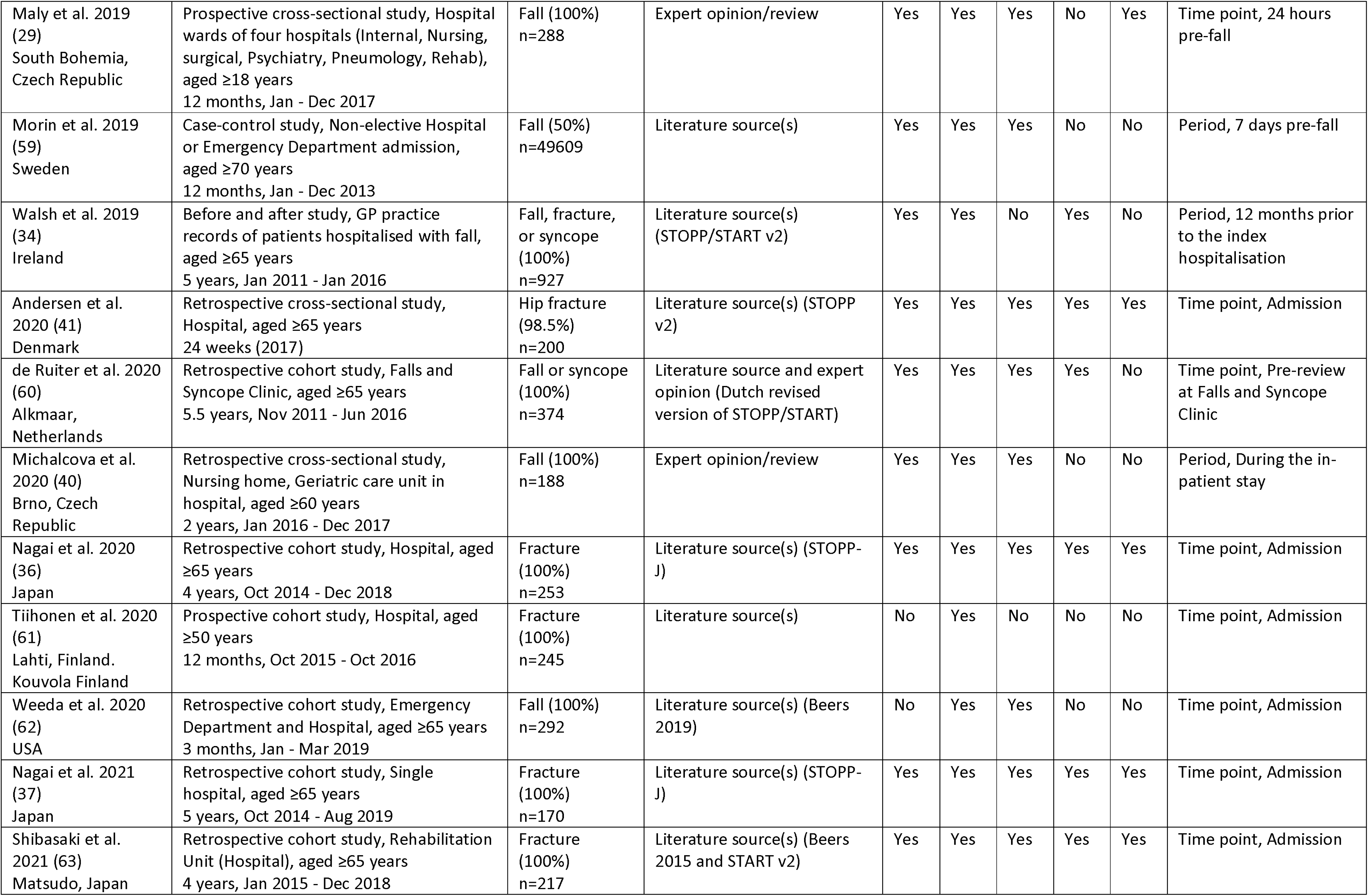
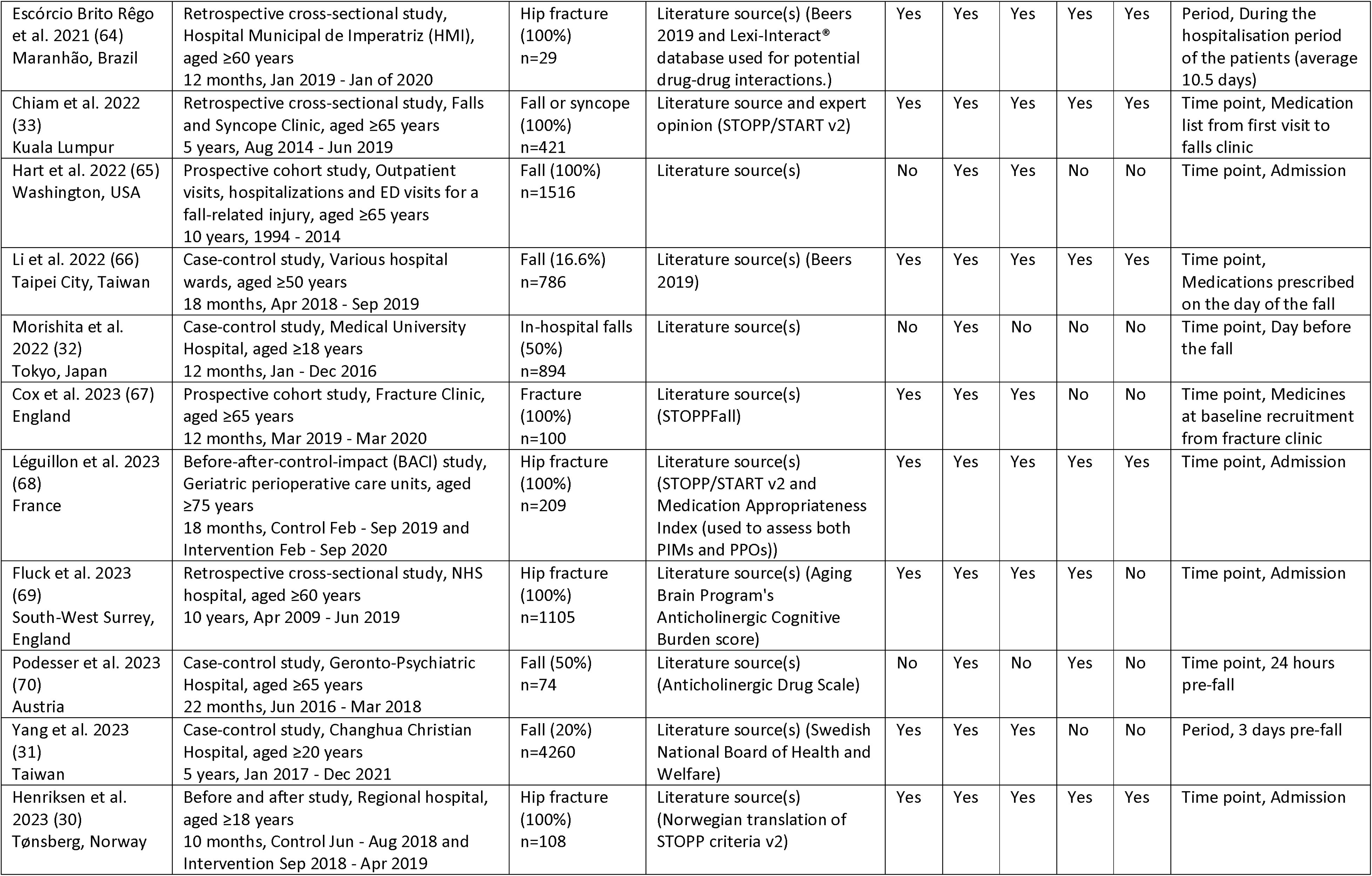
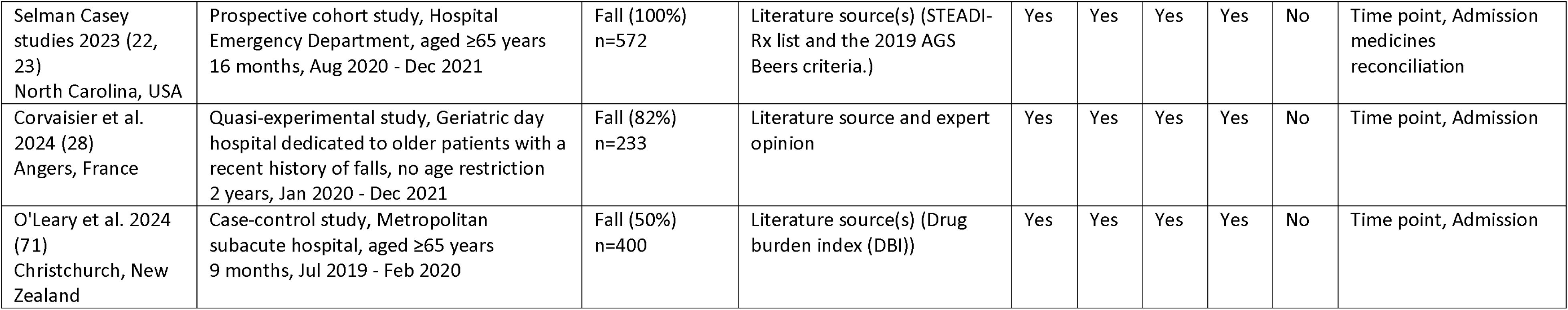
Descriptive characteristics of included studies, listed by year of publication.

| Study, year, location | Study design, setting, time frame | Fall-related inclusion (% with event), sample size | Source of PIP definition (specific tool used) | PIP definition categories |  |  |  |  | Time frame of PIP prevalence measurement |
| --- | --- | --- | --- | --- | --- | --- | --- | --- | --- |
|  |  |  |  | FRID CVD | FRID Psy | FRID Oth | PIP Fall | PIP Oth |  |
| van der Velde et al. 2007 (42)<br>Rotterdam, Netherlands | Prospective cohort study, Geriatric Outpatient Clinic, ≥65 years<br>19 months, Apr 2003 - Nov 2004 | Fall (100%)<br>n=139 | Literature source(s) | Yes | Yes | Yes | No | No | Time point, Admission to Geriatric Outpatient Department |
| Sjöberg et al. 2010 (43)<br>Göteborg, Sweden | Prospective cohort study, Hospital, ≥65 years<br>8 months, Mar - Oct 2008 | Hip fracture (100%)<br>n=100 | Literature source(s) | Yes | Yes | Yes | Yes | No | Time point, Admission |
| Kragh et al. 2011 (44)<br>Sweden | Retrospective cohort study, Hospital ≥60 years<br>12 months, 2006 | Hip fracture (100%)<br>n=2,043 | Literature source(s) | Yes | Yes | Yes | Yes | No | Period, 6 months pre-hip fracture |
| Freeland et al. 2012 (45)<br>South Carolina, USA | Retrospective cross-sectional study, Outpatient internal medicine clinic, ≥65 years<br>13 months | Fall (100%)<br>n=116 | Literature source(s) (Beers 2003) | Yes | Yes | Yes | No | No | Time point, At time of the fall |
| Sjöberg Lönnbro Belfrage studies 2013 (24-26)<br>Göteborg, Sweden | Randomised controlled trial, Department of Geriatrics (Hospital), aged ≥65 years<br>6 months, Apr - Sep 2009 | Hip fracture (100%)<br>n=132 | Literature source(s) (STOPP/START v1 and FRID list from Swedish National Board of Health and Welfare) | Yes | Yes | Yes | Yes | Yes | Time point, Admission |
| Bennett et al. 2014 (46)<br>Sydney, Australia | Prospective cohort study, Tertiary Hospital (aged care, orthopaedic, general medicine department), aged ≥60 years<br>10 months, Jun 2012 - Mar 2013 | Fall (100%)<br>n=204 | Literature source(s) | Yes | Yes | Yes | No | No | Time point, Admission |
| Hohmann et al. 2014 (47)<br>California, USA | Prospective cohort study, Trauma Center (level 2), aged ≥65 years<br>15 months, Aug 2009 - Oct 2010 | Fall (100%)<br>n=112 | Literature source(s) (Beers 2003 and 2012, STOPP/START v1) | Yes | Yes | Yes | Yes | No | Time point, Admission |
| McMahon et al. 2014 (48)<br>Ireland | Before and after study, Hospital Emergency Department, aged ≥70 years<br>4 years, 2007 - 2010 | Fall (100%)<br>n=1016 | Literature source(s) (STOPP v1 and Beers 2012) | Yes | Yes | Yes | Yes | Yes | Period, 12 months pre-fall |
| Rojas-Fernandez et al. 2015 (39)<br>Ontario, Canada | Retrospective cross-sectional study, Retirement Facility, aged ≥65 years<br>12 months, Jul 2009 - Jul 2010 | Fall (100%)<br>n=105 | Literature source(s) | Yes | Yes | Yes | Yes | No | Time point, List of current medications at time of fall |
| Munson et al. 2016 (27)<br>USA | Retrospective cohort study, Community dwelling individuals, no age restriction<br>5 years, 2007 - 2011 | Fracture (100%)<br>n=168133 | Literature source(s) | Yes | Yes | Yes | Yes | No | Period, 120 days pre-fall |
| De Winter et al. 2016 (49)<br>Flanders, Belgium | Case-control study, Traumatology ward (Hospital), aged ≥75 years<br>7 months, Nov 2012 - Jun 2013 | Fall-related fracture (33.5%)<br>n=182 | Literature source and expert opinion | Yes | Yes | Yes | Yes | No | Time point, Admission medicines reconciliation |
| Bambina et al. 2017 (50)<br>France | Prospective cross-sectional study, Hospital (orthopaedic and trauma department), aged ≥65 years<br>6 months, Nov 2015 - Apr 2016 | Fall-related fracture (100%)<br>n=89 | Not stated | Yes | Yes | Yes | No | No | Time point, Admission |
| Blachman et al. 2017 (51)<br>New York, USA | Retrospective cross-sectional study, Hospital, aged ≥65 years<br>1 year, 2012 | Fall (100%)<br>n=328 | Literature source(s) | No | Yes | Yes | No | No | Time point, 24 hours pre-fall |
| Komagamine et al. 2017 (52)<br>Japan | Retrospective cohort study, Hospital, aged ≥65 years<br>2 years, Jan 2015 - Dec 2016 | Hip fracture (100%)<br>n=164 | Literature source and expert opinion (Beers 2015) | Yes | Yes | Yes | Yes | No | Time point, Admission |
| Marvin et al. 2017 (53)<br>London, England | Retrospective cohort study, Hospital Emergency Department, aged ≥70 years<br>2 months, Feb - Mar 2015 | Fall (100%)<br>n=100 | Literature source and expert opinion (STOPP v2) | Yes | Yes | Yes | No | No | Time point, Admission |
| Ryan-Atwood et al. 2017 (54)<br>Victoria, Australia | Case-control study, Specialist Trauma Centre, aged ≥65 years<br>2 years, Jul 2013 - Jun 2015 | Fall (73.8%)<br>n=646 | Literature source(s) | Yes | Yes | Yes | No | No | Time point, Admission medicines reconciliation |
| Beunza-Sola et al. 2018 (55)<br>Navaree, Spain | Prospective cohort study, Tertiary level hospital, aged ≥65 years<br>4 months, Mar - Jun 2016 | Fall-related fracture (100%)<br>n=252 | Literature source(s) (STOPP v1) | Yes | Yes | Yes | No | No | Time point, Admission |
| Lawson et al. 2018 (56)<br>Texas/Mexico Border Community | Retrospective cross-sectional study, Community dwelling older adults, aged ≥55 years<br>Not reported | Fall (100%)<br>n=99 | Literature source(s) (Beers 2012) | Yes | Yes | Yes | Yes | Yes | Time point, Baseline study data collection, within 1 month of fall |
| Machado-Duque studies 2018 (20, 21)<br>Colombia | Case-control study, Community Pharmacy dispensing records of those with a hip fracture recorded in the General System of Social Security in Health of Colombia, aged ≥65 years<br>1 year, Jan - Dec 2015 | Fracture (33.3%)<br>n=900 | Literature source and expert opinion (Salahudeen extended Anticholinergic Risk Scale) | Yes | Yes | Yes | Yes | No | Period, 30 days prior to index date |
| Early et al. 2019 (57)<br>USA | Case-control study, Inpatient, outpatient, and ambulatory care (Medicare database), aged ≥65 years<br>3 years, Jan 2013 - Sep 2015 | Fall (6.6%)<br>n=1678037 | Literature source(s) | Yes | Yes | Yes | No | No | Period, 90 days pre-fall |
| Gleich et al. 2019 (58)<br>Germany | Retrospective cohort study, Trauma centre (level 1), aged ≥70 years<br>2 months, Jul 2016 - Sep 2016 | Hip or humerus fracture (100%)<br>n=95 | Literature source(s) (STOPP/START v2 and PRISCUS list) | Yes | Yes | Yes | Yes | Yes | Time point, Admission |
| Maly et al. 2019 (29)<br>South Bohemia, Czech Republic | Prospective cross-sectional study, Hospital wards of four hospitals (Internal, Nursing, surgical, Psychiatry, Pneumology, Rehab), aged ≥18 years<br>12 months, Jan - Dec 2017 | Fall (100%)<br>n=288 | Expert opinion/review | Yes | Yes | Yes | No | Yes | Time point, 24 hours pre-fall |
| Morin et al. 2019 (59)<br>Sweden | Case-control study, Non-elective Hospital or Emergency Department admission, aged ≥70 years<br>12 months, Jan - Dec 2013 | Fall (50%)<br>n=49609 | Literature source(s) | Yes | Yes | Yes | No | No | Period, 7 days pre-fall |
| Walsh et al. 2019 (34)<br>Ireland | Before and after study, GP practice records of patients hospitalised with fall, aged ≥65 years<br>5 years, Jan 2011 - Jan 2016 | Fall, fracture, or syncope (100%)<br>n=927 | Literature source(s) (STOPP/START v2) | Yes | Yes | No | Yes | No | Period, 12 months prior to the index hospitalisation |
| Andersen et al. 2020 (41)<br>Denmark | Retrospective cross-sectional study, Hospital, aged ≥65 years<br>24 weeks (2017) | Hip fracture (98.5%)<br>n=200 | Literature source(s) (STOPP v2) | Yes | Yes | Yes | Yes | Yes | Time point, Admission |
| de Ruiter et al. 2020 (60)<br>Alkmaar, Netherlands | Retrospective cohort study, Falls and Syncope Clinic, aged ≥65 years<br>5.5 years, Nov 2011 - Jun 2016 | Fall or syncope (100%)<br>n=374 | Literature source and expert opinion (Dutch revised version of STOPP/START) | Yes | Yes | Yes | Yes | No | Time point, Pre-review at Falls and Syncope Clinic |
| Michalcova et al. 2020 (40)<br>Brno, Czech Republic | Retrospective cross-sectional study, Nursing home, Geriatric care unit in hospital, aged ≥60 years<br>2 years, Jan 2016 - Dec 2017 | Fall (100%)<br>n=188 | Expert opinion/review | Yes | Yes | Yes | No | No | Period, During the in-patient stay |
| Nagai et al. 2020 (36)<br>Japan | Retrospective cohort study, Hospital, aged ≥65 years<br>4 years, Oct 2014 - Dec 2018 | Fracture (100%)<br>n=253 | Literature source(s) (STOPP-J) | Yes | Yes | Yes | Yes | Yes | Time point, Admission |
| Tiihonen et al. 2020 (61)<br>Lahti, Finland. Kouvola Finland | Prospective cohort study, Hospital, aged ≥50 years<br>12 months, Oct 2015 - Oct 2016 | Fracture (100%)<br>n=245 | Literature source(s) | No | Yes | No | No | No | Time point, Admission |
| Weeda et al. 2020 (62)<br>USA | Retrospective cohort study, Emergency Department and Hospital, aged ≥65 years<br>3 months, Jan - Mar 2019 | Fall (100%)<br>n=292 | Literature source(s) (Beers 2019) | No | Yes | Yes | No | No | Time point, Admission |
| Nagai et al. 2021 (37)<br>Japan | Retrospective cohort study, Single hospital, aged ≥65 years<br>5 years, Oct 2014 - Aug 2019 | Fracture (100%)<br>n=170 | Literature source(s) (STOPP-J) | Yes | Yes | Yes | Yes | Yes | Time point, Admission |
| Shibasaki et al. 2021 (63)<br>Matsudo, Japan | Retrospective cohort study, Rehabilitation Unit (Hospital), aged ≥65 years<br>4 years, Jan 2015 - Dec 2018 | Fracture (100%)<br>n=217 | Literature source(s) (Beers 2015 and START v2) | Yes | Yes | Yes | Yes | Yes | Time point, Admission |
| Escórcio Brito Rêgo et al. 2021 (64)<br>Maranhão, Brazil | Retrospective cross-sectional study, Hospital Municipal de Imperatriz (HMI), aged ≥60 years<br>12 months, Jan 2019 - Jan of 2020 | Hip fracture (100%)<br>n=29 | Literature source(s) (Beers 2019 and Lexi-Interact® database used for potential drug-drug interactions.) | Yes | Yes | Yes | Yes | Yes | Period, During the hospitalisation period of the patients (average 10.5 days) |
| Chiam et al. 2022 (33)<br>Kuala Lumpur | Retrospective cross-sectional study, Falls and Syncope Clinic, aged ≥65 years<br>5 years, Aug 2014 - Jun 2019 | Fall or syncope (100%)<br>n=421 | Literature source and expert opinion (STOPP/START v2) | Yes | Yes | Yes | Yes | Yes | Time point, Medication list from first visit to falls clinic |
| Hart et al. 2022 (65)<br>Washington, USA | Prospective cohort study, Outpatient visits, hospitalizations and ED visits for a fall-related injury, aged ≥65 years<br>10 years, 1994 - 2014 | Fall (100%)<br>n=1516 | Literature source(s) | No | Yes | Yes | No | No | Time point, Admission |
| Li et al. 2022 (66)<br>Taipei City, Taiwan | Case-control study, Various hospital wards, aged ≥50 years<br>18 months, Apr 2018 - Sep 2019 | Fall (16.6%)<br>n=786 | Literature source(s) (Beers 2019) | Yes | Yes | Yes | Yes | Yes | Time point, Medications prescribed on the day of the fall |
| Morishita et al. 2022 (32)<br>Tokyo, Japan | Case-control study, Medical University Hospital, aged ≥18 years<br>12 months, Jan - Dec 2016 | In-hospital falls (50%)<br>n=894 | Literature source(s) | No | Yes | No | No | No | Time point, Day before the fall |
| Cox et al. 2023 (67)<br>England | Prospective cohort study, Fracture Clinic, aged ≥65 years<br>12 months, Mar 2019 - Mar 2020 | Fracture (100%)<br>n=100 | Literature source(s) (STOPPFall) | Yes | Yes | Yes | No | No | Time point, Medicines at baseline recruitment from fracture clinic |
| Léguillon et al. 2023 (68)<br>France | Before-after-control-impact (BACI) study, Geriatric perioperative care units, aged ≥75 years<br>18 months, Control Feb - Sep 2019 and Intervention Feb - Sep 2020 | Hip fracture (100%)<br>n=209 | Literature source(s) (STOPP/START v2 and Medication Appropriateness Index (used to assess both PIMs and PPOs)) | Yes | Yes | Yes | Yes | Yes | Time point, Admission |
| Fluck et al. 2023 (69)<br>South-West Surrey, England | Retrospective cross-sectional study, NHS hospital, aged ≥60 years<br>10 years, Apr 2009 - Jun 2019 | Hip fracture (100%)<br>n=1105 | Literature source(s) (Aging Brain Program's Anticholinergic Cognitive Burden score) | Yes | Yes | Yes | Yes | No | Time point, Admission |
| Podesser et al. 2023 (70)<br>Austria | Case-control study, Geronto-Psychiatric Hospital, aged ≥65 years<br>22 months, Jun 2016 - Mar 2018 | Fall (50%)<br>n=74 | Literature source(s) (Anticholinergic Drug Scale) | No | Yes | No | Yes | No | Time point, 24 hours pre-fall |
| Yang et al. 2023 (31)<br>Taiwan | Case-control study, Changhua Christian Hospital, aged ≥20 years<br>5 years, Jan 2017 - Dec 2021 | Fall (20%)<br>n=4260 | Literature source(s) (Swedish National Board of Health and Welfare) | Yes | Yes | Yes | No | No | Period, 3 days pre-fall |
| Henriksen et al. 2023 (30)<br>Tønsberg, Norway | Before and after study, Regional hospital, aged ≥18 years<br>10 months, Control Jun - Aug 2018 and Intervention Sep 2018 - Apr 2019 | Hip fracture (100%)<br>n=108 | Literature source(s) (Norwegian translation of STOPP criteria v2) | Yes | Yes | Yes | Yes | Yes | Time point, Admission |
| Selman Casey studies 2023 (22, 23)<br>North Carolina, USA | Prospective cohort study, Hospital Emergency Department, aged ≥65 years<br>16 months, Aug 2020 - Dec 2021 | Fall (100%)<br>n=572 | Literature source(s) (STEADI-Rx list and the 2019 AGS Beers criteria.) | Yes | Yes | Yes | Yes | No | Time point, Admission<br>medicines<br>reconciliation |
| Corvaisier et al. 2024 (28)<br>Angers, France | Quasi-experimental study, Geriatric day hospital dedicated to older patients with a recent history of falls, no age restriction<br>2 years, Jan 2020 - Dec 2021 | Fall (82%)<br>n=233 | Literature source and expert opinion | Yes | Yes | Yes | Yes | No | Time point, Admission |
| O'Leary et al. 2024 (71)<br>Christchurch, New Zealand | Case-control study, Metropolitan subacute hospital, aged ≥65 years<br>9 months, Jul 2019 - Feb 2020 | Fall (50%)<br>n=400 | Literature source(s) (Drug burden index (DBI)) | Yes | Yes | Yes | Yes | No | Time point, Admission |

The most common age restriction was ≥65 years (n=26); all but six studies required participants to be ≥50 years. Two studies had no age restriction,(27, 28) although one of these included geriatric day hospital patients, and four required patients to be adults (aged ≥18/≥20 years).(29–32) Mean or median age of participants was ≥65 years in all studies, and was ≥80 years in thirty-three studies, while females accounted for most fallers in all studies except five (where between 53.9% and 62.1% of fallers were male).

Twenty-three studies included participants with falls or fall events, twenty studies focused on fractures, while two studies focused on falls and syncope,(30, 33) and one on falls, fractures and syncope.(34) Falls and fractures were defined in different ways across the studies. Twenty-two studies described fall events using different free-text definitions which were developed with reference to literature. Twelve studies defined falls/fractures using International Classification of Diseases (ICD) codes.(35) Nine studies did not report how falls/fractures were defined. One study used the AO/OTA fracture and dislocation classification compendium to define a fracture.(36) One study used the visual SQ method to define vertebral fractures as proposed by Genant et al.(37, 38) One study used the French Society of Geriatrics and Gerontology criteria for serious falls.(28)

Thirty-three studies included inpatients and/or attendees at emergency departments, six studies included attendees of an outpatient clinic, five studies included community-dwelling individuals with a record of a fall across inpatient and outpatient/ambulatory care settings (or unspecified settings), and one study each included individuals from a retirement facility,(39) and from both a nursing home and hospital geriatric care unit.(40)

Numbers of participants ranged from 29 to 1,678,037. In 34 studies, 100% of their participants had a fall, one study of 200 consecutive hip fracture patients identified that 98.5% of participants had a fall (41), while another study in a geriatric day hospital included 82% of participants with a recent fall history.(28) In the ten case-control studies, the percentage of fallers included in the study ranged from 6.6% to 73.8%.

### Definitions of PIP

In assessing potentially inappropriate prescribing among fallers, all studies considered FRIDs (in some cases as part of a validated tool for PIP). Three studies assessed only a single category of FRIDs (psychotropics), four studies assessed two categories of FRIDs (three studies assessed psychotropic and other categories, and one study assessed psychotropic and cardiovascular categories), while the remaining thirty-nine studies assessed all three categories. Overall, 46, 42 and 40 studies assessed psychotropic, other, and cardiovascular FRIDs respectively. Twenty-nine studies also assessed other forms of PIP, of which twenty-eight assessed PIP relevant to falls, and fourteen studies assessed non-fall relevant PIP.

Thirty-six studies cited literature source(s) for their PIP definition (including FRIDs), seven cited literature source(s) and expert opinion, two studies referred to expert opinion/review alone, and one did not state the source. Twenty-six studies reported the use of a validated tool for defining PIP. Seventeen studies used a single tool, most often the Beers criteria (five studies) or the STOPP criteria (six studies), and one study each used the STOPPFall criteria, Drug Burden Index, Anticholinergic Cognitive Burden score, Anticholinergic Drug Scale, the Salahudeen extended Anticholinergic Rating Scale, and the Swedish National Board of Health and Welfare indicators. Eleven studies used combinations of tools, most often STOPP/START (n=3), with one study each using other combinations of tools (STOPP/START and Beers criteria, STOPP/START and the PRISCUS list, STOPP/START and Sweden’s FRID list, STOPP/START and Medication Appropriateness Index, STOPP and Beers, START and Beers, Beers and Lexi-Interact®, and Beers and the STEADI-Rx list).

Thirty-six studies assessed prevalence of PIP at a time point, most often reported as at admission (twenty-five studies) followed by at the time of or on the day of the fall (seven studies), while ten studies assessed PIP prevalence over a time period, ranging from three days to twelve months pre-fall.

### Overall prevalence of PIP

A measure of the prevalence of PIP was reported by all included studies, with the percentage of participants with PIP reported in forty-three studies, and the mean number of PIP occurrences per participant was reported or calculable in twenty-three studies (table 1). Twenty studies reported both measures.

The prevalence ranged from 15% to 99%. Across 317,914 participants in included studies, the pooled prevalence (Figure 2) was estimated at 68.6% (95%CI 66.1%, 71.2%), however there was substantial between-study heterogeneity (I^2^ 99.5%, Cochran’s Q p<0.001). Heterogeneity was examined in sub-group analysis, and this was not explained by any aspect of study design or included participant characteristics (see supplementary table 6).

**Figure 2.**
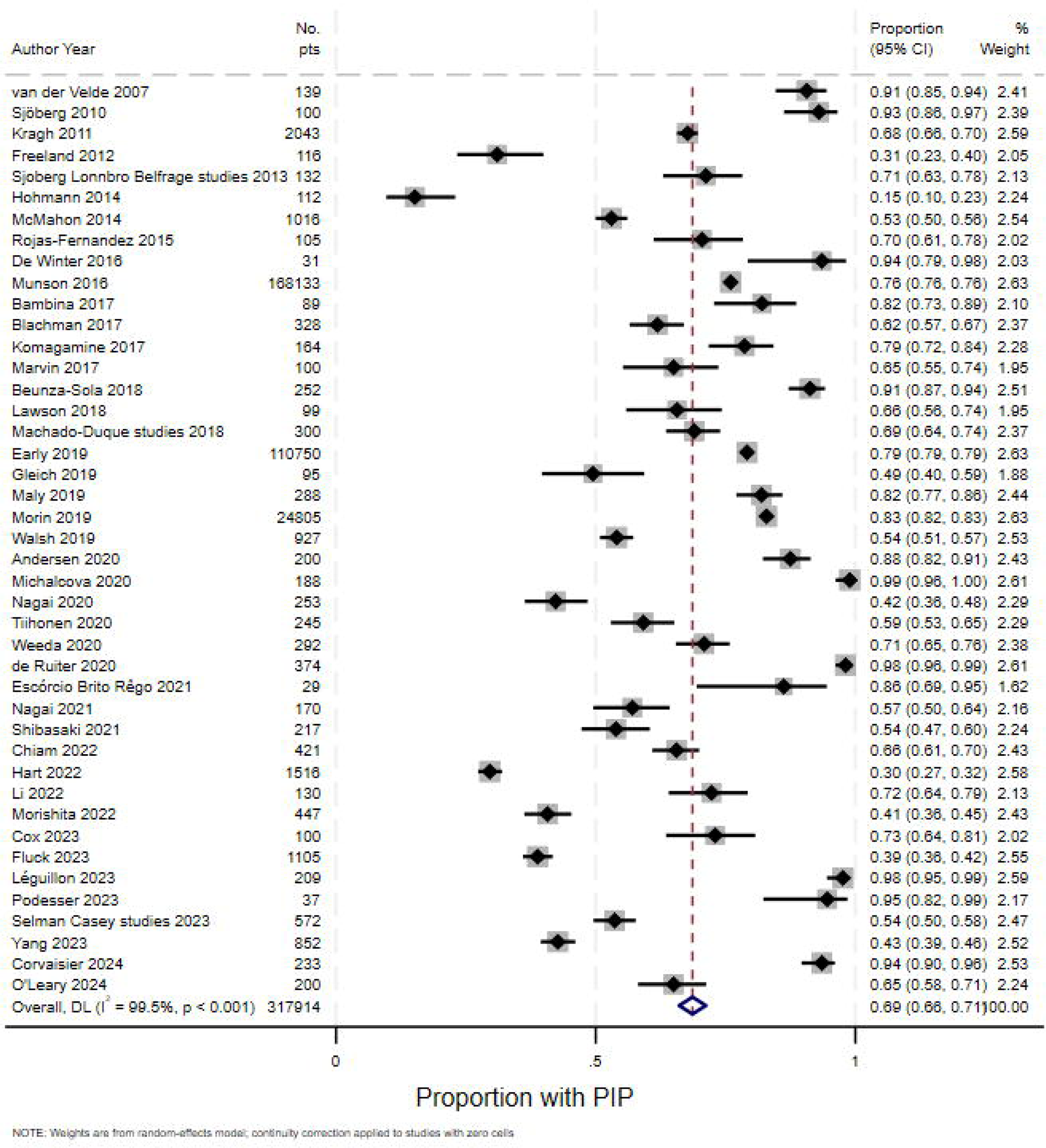
Forest plot for random-effects meta-analysis of studies reporting prevalence of potentially inappropriate prescribing

In the twenty-three studies reporting the mean number of PIP occurrences per participant, this ranged from 0.6 to 5.1. Overall two studies reported means less than 1, nine between 1 and 2, five between 2 and 3, and seven between 3 and 4. The pooled mean (Figure 3) was estimated at 2.21 (95%CI 1.98, 2.45) PIP occurrences per participant, however there was substantial between-study heterogeneity (I^2^ 99.5%, Cochran’s Q p<0.001). Overall prevalence was not explained by study design or included participant characteristics, with the exception of whether PIP was assessed at a time point (mean 2.12, 95%CI 1.65, 2.68) or over a period (mean 2.77, 95%CI 2.38, 3.16), p=0.034 for Cochran’s Q statistic for between-group heterogeneity (see supplementary table 7).

**Figure 3.**
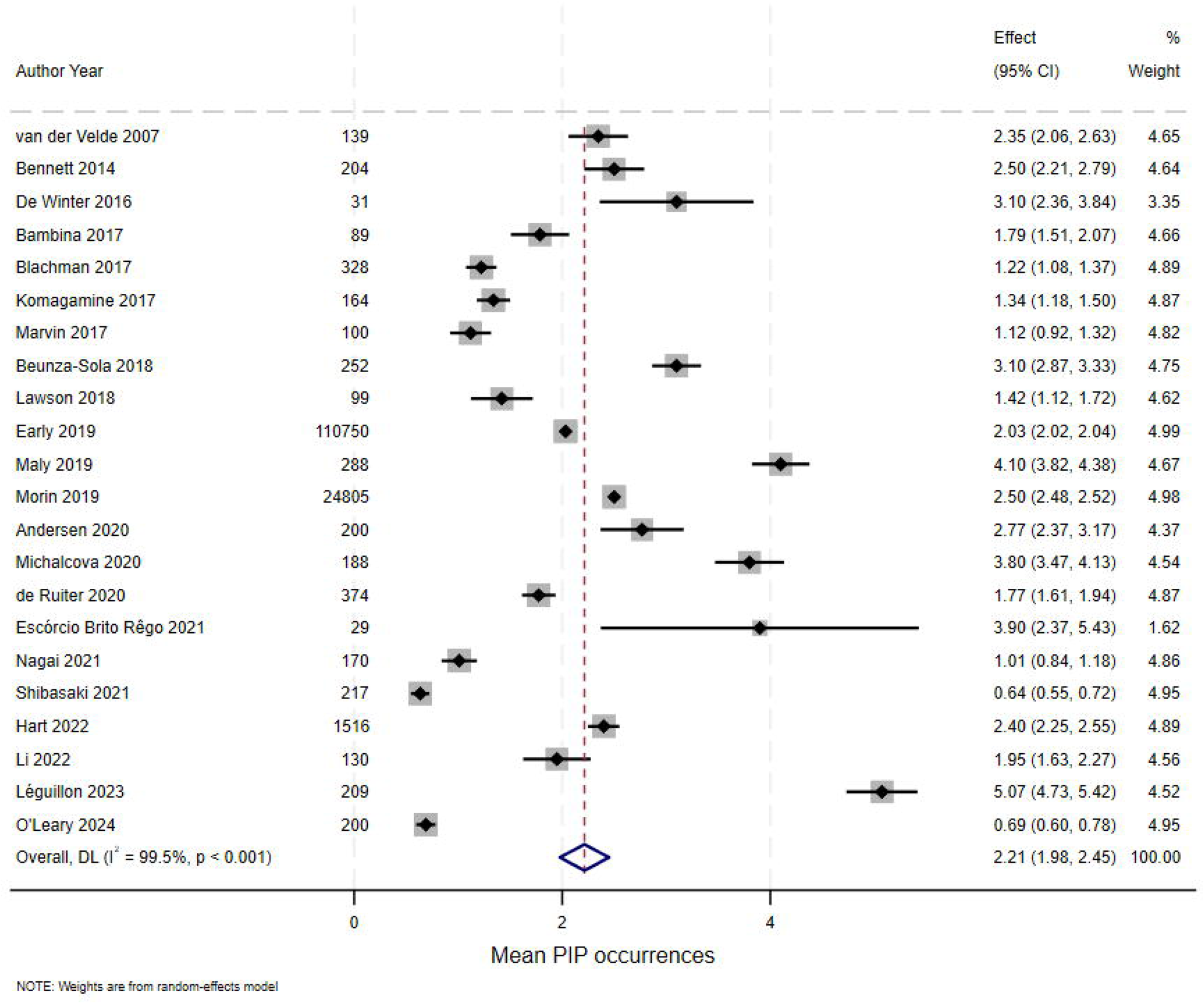
Forest plot for random-effects meta-analysis of studies reporting mean number of potentially inappropriate prescribing occurrences per participant

### Prevalence of individual PIP drugs

Thirty-five studies reported on the prevalence of different drug classes implicated in PIP, and up to the top five most prevalent are reported in Figure 4. Sedative/hypnotic drugs and opioids were reported in 13 studies each, with the percentage of participants prescribed them in the ranges of 3.6-36.5% and 8%-38.1% respectively. The next most frequently reported in twelve studies each were antidepressants (7.5%-56%) and diuretics (12%-60.4%).

**Figure 4.**
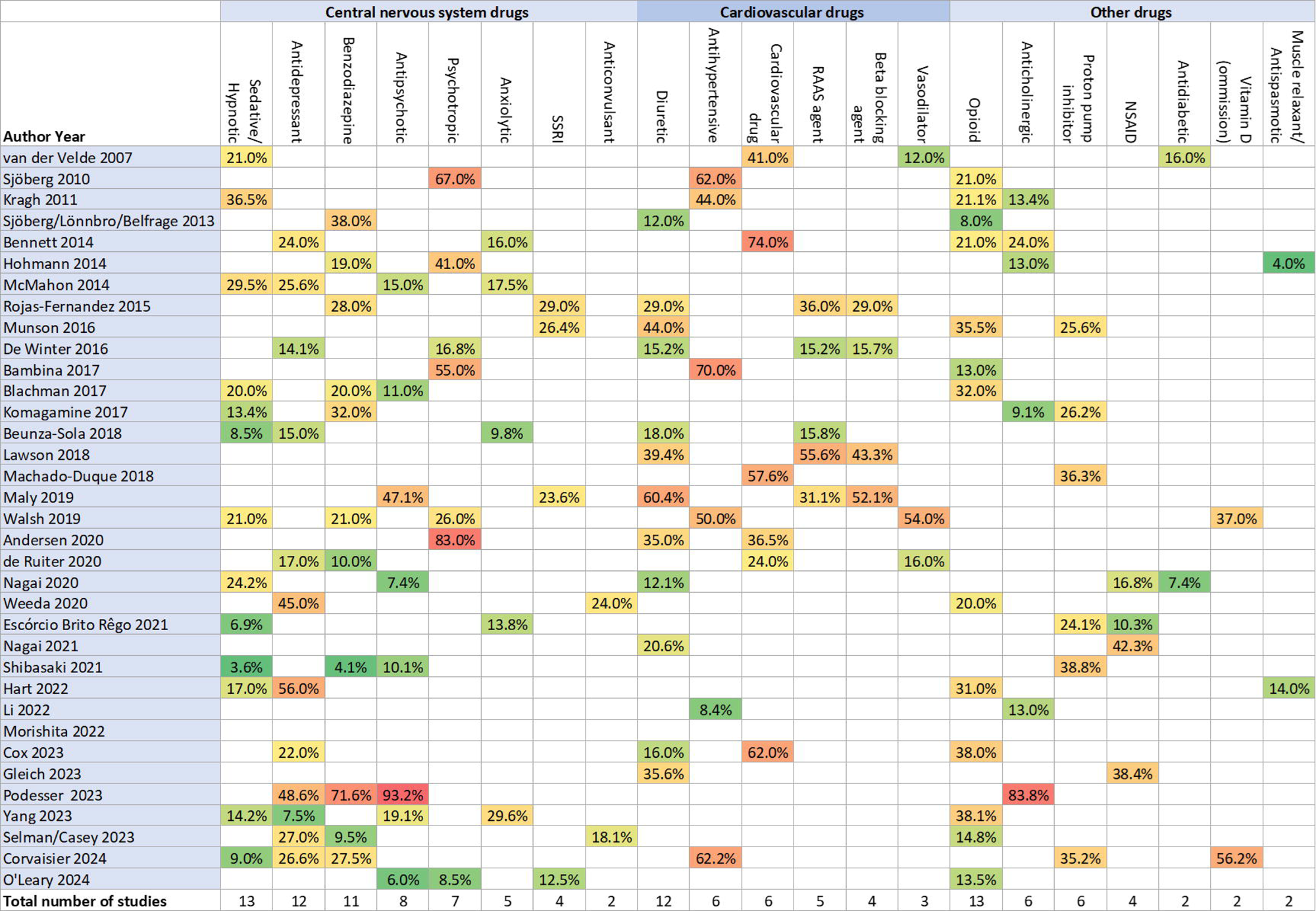
Heat plot of the prevalence of the top 5 drug classes reported as involved in potentially inappropriate prescribing (including falls-risk increasing drugs) *Note: Drug classes only reported among the top 5 classes in fewer than two studies were omitted*.

### Changes in PIP prescribing

Twenty-one studies examined change in prevalence of PIP after the fall (supplementary table 8), with eleven of these measuring changes in prevalence at discharge (and two of these also followed up at six and twelve months). The remaining ten measured changes in prevalence over the following one, three, four, six and/or twelve months. An increase in PIP post-fall was identified in five studies.(24, 34, 44, 55, 62) Three studies identified no change, while a further five identified mixed results (i.e. increases, decreases, and/or no change across different drug classes or follow-up periods). The remaining nine studies identified reductions in PIP post-fall.

## Discussion

Overall, this systematic review of 46 studies identified that PIP is common among fallers internationally. Most studies involved ED/hospitalised patients 65 years and over, all evaluated FRID use while approximately two-thirds also assessed other forms of PIP. The pooled PIP prevalence was 68.6%, with fallers having 2.2 PIP occurrences on average. The most commonly reported drug classes were opioids, sedatives/hypnotics, antidepressants, and diuretics. Fewer than half of studies (n=21) evaluated change in PIP over time, and just under half of these found a reduction in PIP post fall. Few studies examined non-falls-related PIP, which makes it challenging to compare the most prevalent types of PIP among fallers with previous research in general populations, and suggests a need for further research.

Opioids, sedatives/hypnotics, antidepressants, and diuretics were the medication classes most frequently reported as falls-risk increasing or potentially inappropriate in fallers, corresponding to the three major categories of FRIDs. Diuretics have been shown in the literature to be a leading cause of medication induced orthostatic hypotension and volume depletion in older patients,(72) which may explain their high prevalence among fallers. Opioids can cause sedation and cognitive impairment, with pharmacokinetic changes in older adults amplifying these effects and increasing falls risk.(73) Sedative and hypnotic medications such as benzodiazepines are among the most prescribed psychotropic medication, and particularly with chronic use leading to dependence and tolerance, they can cause sedation, impaired balance and potentially cognitive impairment, all risk factors for falls.(74) The broad clinical domains of these medications underlines the importance of a holistic assessment of prescribing appropriateness among fallers, especially those with multiple chronic conditions.

Deprescribing long-term medications can be difficult and may explain why medications deemed to be potentially inappropriate may be continued. Various deprescribing guidelines are available, including for benzodiazepines and opioids,(75, 76) while a diuretics guideline is in development.(77) These provide evidence based recommendations to support decision-making, covering how to identify when and how to reduce or stop medications which are no longer necessary or where potential risks outweigh benefits. An adverse event such as a fall may provide strong support to consider deprescribing, weighed against potentially beneficial effects of the medication.(78) It may be clinically appropriate to continue some potentially inappropriate prescriptions after a fall where the long-term benefits outweigh the anticipated harms, and so some level of post-fall FRID use may be appropriate. Evidence to date on the effect of deprescribing interventions for falls prevention has been mixed,(79, 80) and further robust evaluations of the impact of such interventions as part of multifactorial strategies among patients with falls would be beneficial.

Notably there was substantial between-study heterogeneity both in the proportion of people with falls who had PIP, and mean PIP occurrences. For mean PIP occurrences, this was partly explained by whether PIP was measured at a time period or over a period, indicating a time point may not capture the full complexity of medication use.(81) However, substantial heterogeneity remained among studies, and this could not be explained by reported study-level characteristics. Studies used a variety of different definitions for PIP and reported these differently (e.g. total prevalence or prevalence per validated tool). Even among studies using the same tool, such as STOPP/START or Beers criteria, different versions of these or adaptations to the local context (e.g. due to lacking the data required for application or medications not available in a jurisdiction) may contribute to heterogeneity in prevalence estimates. Future studies should ensure that any such adaptations are clearly reported as some studies did not clearly describe which criteria were applied or omitted. Considering that there are a multitude of factors that contribute to PIP, it is likely that other characteristics not measured or reported at both study- and participant-level e.g. prevalence of particular conditions or number of medicines, may further explain heterogeneity.

### Strengths and limitations

A strength of this review is the inclusive approach for defining falls and related-events, populations and settings, and potentially inappropriate prescribing, yielding a comprehensive synthesis of research in this area, which complements other more focused evidence synthesis such as the review of falls-related injuries, which included only 14 studies, all of which were included in this review.(12) The review protocol was pre-registered and followed methodological guidance for systematic reviews of prevalence studies. Limitations include the focus on peer-reviewed literature, given the likely low contribution of grey literature sources to the topic, however potentially relevant research may be omitted. Heterogeneity in prevalence was high, which may reflect differences in study design, populations, drug classes used to define PIP/FRIDs and other aspects not examined in stratified analyses.(18) Within the review timeframe, it was not feasible to contact study authors to obtain information not reported. For example heterogeneity in how drug and drug class prevalence was reported across studies impeded further statistical analysis for specific drugs/classes.

### Implications

Future studies on this topic should adopt a standardised approach to recording and reporting medication use at the individual drug and drug class levels, which would enhance the evidence base for targeted approaches to address FRIDs among fallers. Similarly, more comprehensive assessment of not just FRIDs, but also other forms of PIP among fallers should be considered in further research, including a full description of the criteria applied. Further research is also needed to determine the extent to which medication review and optimisation occurs after a fall, as fewer than half of included studies reported on this.

Depending on the healthcare context, a falls admission may provide an opportunity to review and optimise medicines use in general, for example, where a fall-related admission triggers a comprehensive geriatric assessment. The recent World Guidelines for Fall Prevention and Management recommend medication review and appropriate deprescribing of FRIDs as part of multifactorial falls prevention.(10) However a recent systematic review on the effectiveness of medication review and deprescribing interventions as a single intervention in falls prevention identified wide heterogeneity in interventions and did not identify a significant effect, (although this was among all populations, not specifically people with an existing fall).(80) Likelihood of benefit may be increased by focusing on individuals taking FRIDs with strong evidence of an impact on falls or other factors which predict falls risk.(82, 83)

## Conclusion

The high prevalence among fallers of potentially inappropriate prescribing, including FRID use, identified in this review suggests significant scope for medicines optimisation in this group. This could focus on falls risk reduction and improving prescribing appropriateness more generally, however the evidence that this occurs routinely is limited and mixed. Improved targeting of deprescribing interventions to address key FRID classes, as part of multifactorial falls prevention strategies may ultimately reduce future falls and improve patient outcomes.

Declarations of interest

All authors have no interests to declare

## Funding

No specific funding was received for this study. JGL is supported by funding from the Wellcome Trust (DIAMOND programme, grant number 227348/Z/23/Z).

## Data Availability

All extracted data are contained within the manuscript supplementary materials

## Acknowledgements

TOR was employed by AstraZeneca at the time of manuscript submission. KI is supported by the National Institute for Health and Care Research Applied Research Collaboration Wessex. The views expressed in this publication are those of the authors and not necessarily those of the National Institute for Health and Care Research or the Department of Health and Social Care.

## Notes

### Competing Interest Statement

The authors have declared no competing interest.

