## Supplementary for "Potentially inappropriate prescribing and falls-risk increasing drugs in people who have experienced a fall; a systematic review and meta-analysis"

### Supplementary materials

Supplementary table 1. Search strategy – Medline (ovid)

| 1 | exp Inappropriate Prescribing/ | 4,538 |
| --- | --- | --- |
| 2 | exp Potentially Inappropriate Medication List/ | 966 |
| 3 | (falls-risk drug* or falls risk drug* or fall-risk drug* or fall risk drug* or fall-risk increasing drug* or fall risk increasing drug* or falls-risk increasing drug* or falls risk increasing drug* or inappropriate medic* or inappropriate prescr* or appropriate prescr* or appropriate medic* or inappropriate drug* or appropriate drug*) | 15,330 |
| 4 | ((inappropriat* or appropriat* or suboptim* or "sub‐optim*" or unnecessary) adj1 (medicine* or medicat* or prescrib* or prescription* or drug*)) | 14,839 |
| 5 | (stopp or beers or medication appropriateness index or screening tool of older persons prescriptions or screening tool to alert to right treatment or "stopp/start" or stopp start) or (((beer? or shan? or mcleod?) adj3 criter*) or "fit for the aged" or ((forta or rasp or priscus) adj3 (criter* or list? or instrument))) | 4,465 |
| 6 | 1 or 2 or 3 or 4 or 5 | 21,183 |
| 7 | exp Accidental Falls/ | 27,923 |
| 8 | exp Fractures, Bone/ | 206,778 |
| 9 | (fall? or fell or falling or fallen or faller or stumble* or stumbling or stumbles or slip or slips or slipping or slipped or trip or tripped or fractur*) | 652,454 |
| 10 | 7 or 8 or 9 | 654,818 |
| 11 | 6 and 10 | 998 |

Supplementary table 2. Search strategy – Embase

| 1 | ('inappropriate prescribing'/exp OR 'inappropriate prescribing') AND [embase]/lim | 8,222 |
| --- | --- | --- |
| 2 | (‘falls-risk drug*’ OR ‘falls risk drug*’ OR ‘fall-risk drug*’ OR ‘fall risk drug*’ OR ‘fall-risk increasing drug*’ OR ‘fall risk increasing drug*’ OR ‘falls-risk increasing drug*’ OR ‘falls risk increasing drug*’ OR ‘inappropriate medic*’ OR ‘inappropriate prescr*’ OR ‘appropriate prescr*’ OR ‘appropriate medic*’ OR ‘inappropriate drug*’ OR ‘appropriate drug*’) AND [embase]/lim | 17,529 |
| 3 | ((inappropriat* OR appropriat* OR suboptim* OR ‘sub‐optim*’ OR unnecessary) NEAR/1 (medicine* OR medicat* OR prescrib* OR prescription* OR drug*)) AND [embase]/lim | 17,908 |
| 4 | (stopp OR beers OR ‘medication appropriateness index’ OR ‘screening tool of older persons prescriptions’ OR ‘screening tool to alert to right treatment’ OR ‘stopp/start’ OR ‘stopp start’) OR (((beer? OR shan? OR mcleod?) NEAR/3 criter*) OR ‘fit for the aged’ OR ((forta OR rasp OR priscus) NEAR/3 (criter* OR list? OR instrument))) AND [embase]/lim | 8,097 |
| 5 | #1 OR #2 OR #3 OR #4 | 30,539 |
| 6 | ('falls'/exp OR 'falls') AND [embase]/lim | 88,652 |
| 7 | ('fracture'/exp OR 'fracture') AND [embase]/lim | 352,678 |
| 8 | (fall? OR fell OR falling OR fallen OR faller OR stumble* OR stumbling OR stumbles OR slip OR slips OR slipping OR slipped OR trip OR tripped OR fractur*) AND [embase]/lim | 598,205 |
| 9 | #6 OR #7 OR #8 | 606,076 |
| 10 | #5 AND #9 | 1,740 |

Supplementary table 3. Search strategy – CINAHL PLUS (EBSCOhost)

| 1 | (MH "Inappropriate Prescribing") | 3,489 |
| --- | --- | --- |
| 2 | (“falls-risk drug*” OR “falls risk drug*” OR “fall-risk drug*” OR “fall risk drug*” OR “fall-risk increasing drug*” OR “fall risk increasing drug*” OR “falls-risk increasing drug*” OR “falls risk increasing drug*” OR “inappropriate medic*” OR “inappropriate prescr*” OR “appropriate prescr*” OR “appropriate medic*” OR “inappropriate drug*” OR “appropriate drug*”) | 7,110 |
| 3 | ((inappropriat* OR appropriat* OR suboptim* OR "sub‐optim*" OR unnecessary) N1 (medicine* OR medicat* OR prescrib* OR prescription* OR drug*)) | 8,770 |
| 4 | (stopp OR beers OR “medication appropriateness index” OR “screening tool of older persons prescriptions” OR “screening tool to alert to right treatment” OR “stopp/start” OR “stopp start”) OR (((beer? OR shan? OR mcleod?) N3 criter*) OR “fit for the aged” OR ((forta OR rasp OR priscus) N3 (criter* OR list? OR instrument))) | 4,861 |
| 5 | S1 OR S2 OR S3 OR S4 | 13,806 |
| 6 | (MH "Accidental Falls") | 26,321 |
| 7 | (MH "Fractures") | 21,730 |
| 8 | (fall* OR fell OR falling OR fallen OR faller OR stumble* OR stumbling OR stumbles OR slip OR slips OR slipping OR slipped OR trip OR tripped OR fractur*) | 197,105 |
| 9 | S6 OR S7 OR S8 | 197,105 |
| 10 | S5 AND S9 | 727 |

Google scholar: inappropriate prescribing falls-risk increasing drugs falls fractures (keywords, top 200 results)

Supplementary table 4. Search strategy – Google Scholar)

Searched via Harzing’s Publish or Perish

Terms: inappropriate prescribing falls-risk increasing drugs falls fractures

Restricted to keywords

Restrict to top 200 results by relevance

Supplementary table 5: Quality assessment using Joanna Briggs Institute Prevalence Critical Appraisal tool

| **Author** | 1. Was the sample frame appropriate to address the target population? | 2. Were study participants sampled in an appropriate way? | 3. Was the sample size adequate? | 4. Were the study subjects and the setting described in detail? | 5. Was the data analysis conducted with sufficient coverage of the identified sample? | 6. Were valid methods used for the identification of the condition? | 7. Was the condition measured in a standard, reliable way for all participants? | 8. Was there appropriate statistical analysis? | 9. Was the response rate adequate, and if Not, was the low response rate managed appropriately? |
| --- | --- | --- | --- | --- | --- | --- | --- | --- | --- |
| van der Velde et al. 2007 | Yes | Yes | No | Yes | Yes | Yes | Yes | Yes | Not applicable |
| Sjöberg et al. 2010 | Yes | Yes | No | Yes | Yes | Yes | Yes | Yes | Not applicable |
| Kragh et al. 2011 | Yes | Yes | Yes | Yes | Yes | Yes | Yes | Yes | Not applicable |
| Freeland et al. 2012 | Yes | Yes | No | No | No | Yes | No | Yes | Not applicable |
| Sjöberg Lönnbro Belfrage studies 2013 | Yes | Yes | No | No | Yes | Yes | Yes | Yes | Not applicable |
| Bennett et al. 2014 | Yes | Yes | No | No | Yes | Yes | Yes | Yes | Not applicable |
| Hohmann et al. 2014 | Yes | Yes | No | No | Yes | No | Yes | Yes | Not applicable |
| McMahon et al. 2014 | Yes | Yes | Yes | Yes | Yes | Yes | Yes | Yes | Not applicable |
| Rojas-Fernandez et al. 2015 | Yes | Yes | No | Yes | Yes | Yes | Yes | Unclear | Not applicable |
| Munson et al. 2016 | Yes | Yes | Yes | Yes | Yes | Yes | Yes | Yes | Not applicable |
| De Winter et al. 2016 | Unclear | Yes | No | Yes | Yes | Yes | Yes | Yes | Not applicable |
| Bambina et al. 2017 | Yes | Yes | No | No | Unclear | Unclear | Unclear | No | Not applicable |
| Blachman et al. 2017 | Yes | Yes | Yes | No | No | Yes | Yes | No | Not applicable |
| Komagamine et al. 2017 | Yes | No | No | Yes | Yes | Yes | Yes | Yes | Not applicable |
| Marvin et al. 2017 | Unclear | Yes | No | Yes | Unclear | Unclear | Unclear | No | Not applicable |
| Ryan-Atwood et al. 2017 | Yes | Yes | Yes | Yes | Yes | Yes | Yes | Yes | Not applicable |
| Beunza-Sola et al. 2018 | Yes | Yes | Yes | Yes | Yes | Yes | Yes | Yes | Not applicable |
| Lawson et al. 2018 | No | Unclear | No | No | Yes | Unclear | Yes | Unclear | Not applicable |
| Machado-Duque studies 2018 | Yes | Yes | Yes | Yes | Yes | Yes | Yes | Yes | Not applicable |
| Early et al. 2019 | Yes | Yes | Yes | Yes | Yes | Yes | Yes | Yes | Not applicable |
| Gleich et al. 2019 | Yes | Yes | No | Yes | Yes | Yes | Yes | Yes | Not applicable |
| Maly et al. 2019 | Yes | Yes | Yes | Unclear | Yes | Yes | Yes | Unclear | Not applicable |
| Morin et al. 2019 | Yes | Yes | Yes | Yes | Yes | Yes | Yes | Yes | Not applicable |
| Walsh et al. 2019 | Yes | Yes | Yes | Yes | Yes | Yes | Yes | Yes | Not applicable |
| Andersen et al. 2020 | Yes | Yes | No | Yes | Yes | Yes | Yes | Yes | Not applicable |
| de Ruiter et al. 2020 | Yes | Yes | Yes | Yes | Yes | Unclear | Yes | Yes | Not applicable |
| Michalcova et al. 2020 | Yes | Yes | No | Yes | Yes | Yes | Yes | No | Not applicable |
| Nagai et al. 2020 | Yes | Yes | Yes | Yes | Yes | Yes | Yes | Yes | Not applicable |
| Tiihonen et al. 2020 | Yes | Yes | Yes | Yes | Yes | Yes | Yes | Yes | Not applicable |
| Weeda et al. 2020 | Yes | Yes | Yes | No | Unclear | Yes | Yes | Yes | Not applicable |
| Nagai et al. 2021 | Yes | Yes | No | Yes | Yes | Yes | Yes | Yes | Not applicable |
| Shibasaki et al. 2021 | Yes | Yes | Yes | Yes | Yes | Yes | Yes | Yes | Not applicable |
| Escórcio Brito Rêgo et al. 2021 | Yes | Yes | No | Yes | Yes | Yes | Yes | Yes | Not applicable |
| Chiam et al. 2022 | Yes | Yes | Yes | Yes | Yes | Yes | Yes | Yes | Not applicable |
| Hart et al. 2022 | Yes | Yes | Yes | No | Yes | Yes | Yes | Yes | Not applicable |
| Li et al. 2022 | Unclear | Yes | Yes | Yes | Yes | Yes | Yes | Yes | Not applicable |
| Morishita et al. 2022 | Yes | Yes | Yes | Yes | Yes | Yes | Yes | Yes | Not applicable |
| Cox et al. 2023 | Yes | Yes | No | Yes | Yes | Yes | Yes | Yes | Not applicable |
| Léguillon et al. 2023 | Yes | Yes | Yes | Yes | Yes | Yes | Unclear | Yes | Not applicable |
| Fluck et al. 2023 | Yes | Yes | Yes | Yes | Yes | Yes | Yes | Yes | Not applicable |
| Podesser et al. 2023 | Yes | Yes | No | Yes | Unclear | Yes | Yes | Yes | Not applicable |
| Yang et al. 2023 | Yes | Yes | Yes | Yes | Yes | Yes | Yes | Yes | Not applicable |
| Henriksen et al. 2023 | Yes | Unclear | No | Yes | Yes | Yes | Yes | Yes | Not applicable |
| Selman Casey studies 2023 | Yes | No | Yes | Yes | Yes | Yes | Yes | Yes | Not applicable |
| Corvaisier et al. 2024 | Yes | Yes | Yes | Yes | Yes | Yes | Yes | Yes | Not applicable |
| O'Leary et al. 2024 | Yes | Yes | Yes | Yes | Yes | Yes | Yes | Yes | Not applicable |

Supplementary table 6. Stratified meta-analysis of PIP prevalence by study characteristics

|  | Proportion | [95% Conf. | Interval] | % Weight | Cochran's Q statistic | df | p-value | I2 statistic | Between group Cochran's Q and p value | |
| --- | --- | --- | --- | --- | --- | --- | --- | --- | --- | --- |
| Included ED/hospitalised participants | | |  |  |  |  |  |  |  |  |
| No | 0.695 | 0.578 | 0.811 | 25.63 | 2857.54 | 10 | <0.001 | 0.997 | 0.04 | 0.834 |
| Yes | 0.682 | 0.644 | 0.719 | 74.37 | 4382.64 | 31 | <0.001 | 0.993 |  |  |
| Included participants with fractures | | |  |  |  |  |  |  |  |  |
| No | 0.664 | 0.616 | 0.713 | 54.36 | 5317.99 | 22 | <0.001 | 99.60% | 1.25 | 0.263 |
| Yes | 0.71 | 0.646 | 0.775 | 45.64 | 1716.4 | 19 | <0.001 | 98.90% |  |  |
| Measurement of prevalence | | |  |  |  |  |  |  |  |  |
| Time point | 0.677 | 0.584 | 0.771 | 75.32 | 5064.02 | 32 | <0.001 | 99.40% | 0.32 | 0.573 |
| Period | 0.706 | 0.673 | 0.739 | 24.68 | 2668.38 | 9 | <0.001 | 99.70% |  |  |
| Number of PIP categories assessed | | |  |  |  |  |  |  |  |  |
| 1-2 categories | 0.594 | 0.405 | 0.782 | 14.22 | 507.75 | 5 | <0.001 | 0.99 | 2.39 | 0.496 |
| 3 categories | 0.728 | 0.682 | 0.774 | 25.98 | 1773.24 | 10 | <0.001 | 0.994 |  |  |
| 4 categories | 0.711 | 0.628 | 0.794 | 33.41 | 2343.47 | 13 | <0.001 | 0.994 |  |  |
| 5 categories | 0.668 | 0.536 | 0.8 | 26.39 | 897.59 | 11 | <0.001 | 0.988 |  |  |
| Number of FRID categories assessment | | | |  |  |  |  |  |  |  |
| 1 category | 0.647 | 0.35 | 0.945 | 6.89 | 151.85 | 2 | <0.001 | 0.987 | 3.09 | 0.214 |
| 2 categories | 0.54 | 0.348 | 0.733 | 9.86 | 334.8 | 3 | <0.001 | 0.991 |  |  |
| 3 categories | 0.71 | 0.685 | 0.734 | 83.25 | 5493.88 | 35 | <0.001 | 0.994 |  |  |
| Assessed non-fall-related PIP | | |  |  |  |  |  |  |  |  |
| No | 0.688 | 0.659 | 0.718 | 71.16 | 6855.08 | 29 | <0.001 | 99.60% | 0.02 | 0.895 |
| Yes | 0.68 | 0.56 | 0.8 | 28.84 | 902.41 | 12 | <0.001 | 98.70% |  |  |

Supplementary table 7. Stratified meta-analysis of mean PIP occurrences by study characteristics

|  | Mean | [95% Conf. | Interval] | % Weight | Cochran's Q statistic | df | p-value | I2 statistic | Between group Cochran's Q and p value | |
| --- | --- | --- | --- | --- | --- | --- | --- | --- | --- | --- |
| Included ED/hospitalised participants | | |  |  |  |  |  |  |  |  |
| No | 1.99 | 1.56 | 2.43 | 19.03 | 53.12 | 3 | <0.001 | 94.40% | 1.14 | 0.286 |
| Yes | 2.27 | 2.00 | 2.54 | 80.97 | 4220.8 | 17 | <0.001 | 99.60% |  |  |
| Included participants with fractures | |  |  |  |  |  |  |  |  |  |
| No | 2.12 | 1.87 | 2.38 | 62.06 | 2633.9 | 12 | <0.001 | 99.50% | 0.61 | 0.436 |
| Yes | 2.47 | 1.64 | 3.30 | 37.94 | 1018.53 | 8 | <0.001 | 99.20% |  |  |
| Measurement of prevalence | |  |  |  |  |  |  |  |  |  |
| Time point | 2.12 | 1.65 | 2.58 | 83.88 | 1925.88 | 17 | <0.001 | 99.10% | 4.51 | 0.034 |
| Period | 2.77 | 2.38 | 3.16 | 16.12 | 1273.46 | 3 | <0.001 | 99.80% |  |  |
| Number of PIP categories assessed | | |  |  |  |  |  |  |  |  |
| 1-2 categories | 1.81 | 0.66 | 2.97 | 9.78 | 120.28 | 1 | <0.001 | 99.20% | 0.99 | 0.804 |
| 3 categories | 2.38 | 2.12 | 2.64 | 38.03 | 1448.99 | 7 | <0.001 | 99.50% |  |  |
| 4 categories | 2.18 | 1.19 | 3.16 | 22.71 | 611.69 | 4 | <0.001 | 99.30% |  |  |
| 5 categories | 2.33 | 1.33 | 3.34 | 29.48 | 736.58 | 6 | <0.001 | 99.20% |  |  |
| Number of FRID categories assessment | | | |  |  |  |  |  |  |  |
| 2 categories | 1.81 | 0.66 | 2.97 | 9.78 | 120.28 | 1 | <0.001 | 99.20% | 0.55 | 0.458 |
| 3 categories | 2.26 | 2.01 | 2.51 | 90.22 | 4127.78 | 19 | <0.001 | 99.50% |  |  |
| Assessed non-fall-related PIP | | |  |  |  |  |  |  |  |  |
| No | 2.08 | 1.84 | 2.32 | 65.85 | 2576.38 | 13 | <0.001 | 99.50% | 0.76 | 0.384 |
| Yes | 2.57 | 1.48 | 3.66 | 34.15 | 1165.7 | 7 | <0.001 | 99.40% |  |  |

Supplementary table 8: Changes in the prevalence of potentially inappropriate prescribing

| **Study, year** | **Baseline time point/period** | **Timeframe for change in prevalence** | **Overall change** | **Description** |
| --- | --- | --- | --- | --- |
| van der Velde et al. 2007 | Admission to Geriatric Outpatient Department | 3-month follow up | Reduction | 75 (59% of those using FRID) people had medications withdrawn in the 3 months after fall. |
| Sjöberg et al. 2010 | Admission | Discharge and 6-month follow up. | Mixed: increase at discharge and return to baseline at 6 months | FRIDs among 93 (93%) at admission, 96 (100%) at discharge,73 (94%) at 6 months |
| Kragh et al. 2011 | 6 months pre-hip fracture | 6 months post-hip fracture | Increase | FRIDs among 67% pre-fracture and post fracture this increased to 97.7% |
| Sjoberg Lonnbro Belfrage studies et al. 2013 | Admission | Discharge, 6-month, 12 month follow up. | Mixed: increase at discharge and return to baseline at 6/12 months | Among intervention group mean +/- SD at admission was 3.08 +/- 2.23 FRIDs, discharge 3.86 +/- 2.06, 6 Months: 3.03 +/- 2.15, and 12 Months: 2.85 +/-2.05. For the control group this was a mean at admission of 3.06 +/- 1.89, discharge: 4.21 +/- 1.99, 6 months: 3.33 +/- 2.32, and 12 Months 3.09 +/- 2.22 |
| Bennett et al. 2014 | Admission | Discharge | No change | From 2.5 +/- 2.1 on admission to 2.5 +/- 1.9 on discharge (overall change in FRID 0 +/- 2.8) |
| McMahon et al. 2014 | 12 months pre-fall | 12 months post-fall | Mixed: no significant change in STOPP/Beers prevalence, significant reductions in long acting benzos, neuroleptics, sig increase in long term PPIs | 53.1% STOPP pre, 53.7% post. Beers 44% to 41.5%, neither significant. Long acting benzo from 10.7% to 8.6% (P=0.0045), Neuroleptics From 17.5% to 14.7% (P=0.022). Long term PPI increasing from 19.3% to 22.5% (P=0.012) |
| Rojas-Fernandez et al. 2015 | List of current medications at time of fall | Over the 6 months post-fall | No change | Review did not reveal any medication regimen changes in the 6-month period after each fall that was related to medications that might have played a role in falls |
| Munson et al. 2016 | 120 days pre-fall | 120 days post fracture | Increase | Increases in drugs that increase fracture risk across all fracture types (hip: 80.6% to 89.3%, wrist 81.8% to 85.0%, shoulder 86.6% to 89.4%) |
| Komagamine et al. 2017 | Admission | Discharge | Reduction | From 124 taking a PIM on admission (79.1%) to 103 taking a PIM on disharge ( 65.2%) |
| Marvin et al. 2017 | Admission | Discharge | Reduction | 38 (38%) of patients had their medications reduced in some way. 39% of the medications changed were fall risk increasing drugs. |
| Beunza-Sola et al. 2018 | Admission | 1 month post admission | Increase | Increase in FRID post fall from mean 3.1 to 3.4 FRIDs |
| Gleich et al. 2019 | Admission | Discharge | Mixed: no change in one group, reduction in the other | For the conventional trauma ward no adjustment of medication during hospital stay was observed, 48.1% (26 of 54 patients) of the patients on the orthogeriatric ward received an adjustment by the geriatrician |
| Walsh et al. 2019 | 12 months prior to the index hospitalisation | 12 months post hospital discharge | Mixed: increase in benzo/z-drug/neuroleptic, also increase in vitamin D | Benzo From 21% to 23% (P=0.06), Z-drugs from 18% to 22% (P<0.01), neuroleptics from 12% to 15% (P<0.01), vasodilators from 54% to 54% (P=0.7), vitamin D from 37% to 52% (P<0.01) |
| de Ruiter et al. 2020 | Pre-review at Falls and Syncope Clinic | Discharge | Reduction | Reduced from 98% PIP at admission to 91% with at least one PIP at discharge (PIM 62%, PPO 72%) |
| Weeda et al. 2020 | Admission | Discharge | Increase | PIM prevalence increased from 207 patients (70.9%) to 230 (78.8%) of patients at discharge (P<0.001). |
| Hart et al. 2022 | Admission | 3, 6, and 12 months post fall-related injury | Reduction | Standardized daily dose at baseline was 2.4, this decreased by 0.38, 0.39 and 0.43 at 3, 6, and 12 months among users of FRIDs at baseline, and rose by 0.18 among non-users of FRIDs, hence an overall reduction. |
| Cox et al. 2023 | Medications at baseline recruitment from fracture clinic | 3 and 6 month follow up | Reduction | Baseline 73% on a FRID, at 3 month follow up 64% on a FRID. and at 6 month follow up 59%. |
| Léguillon et al. 2023 | Admission | Discharge | Reduction | Patients with ≥1 PIPs decreased from 97.6% to 76.1% at discharge. There was also a reduction in mean number of PIPs |
| Henriksen et al. 2023 | Admission | Discharge | Reduction | Change from admission to discharge among intervention group of mean 1.36 PIMs to 1.00, and among control group of mean 1.94 PIMs to 1.83. |
| Selman Casey studies et al. 2023 | Medication reconciliation at admission | 12 months post fall | Reduction | 43.4% of those with a medication change recommend had a modification made |
| O'Leary et al. 2024 | Admission | Discharge | No change | Change in DBI from admission to discharge was a reduction of 0.03, deemed to not be clinically significant |
